# LEAF- 4L6715 enhances oxygenation in patients with acute respiratory distress syndrome (ARDS) due to severe COVID-19: Final results of a phase I/II clinical trial

**DOI:** 10.1101/2022.09.07.22279668

**Authors:** Paul-Michel Mertes, Xavier Delabranche, Pierre Coliat, Anne Roche, Olivier Collange, Manon Voegelin, Alexandre Bernard, Navreet Dhindsa, Zhenghong Hannah Xu, Bolin Geng, Clet Niyikiza, Victor Moyo, Claire Bourban, Pascal Villa, Alexandre Detappe, Xavier Pivot

## Abstract

LEAF-4L6715 is a liposomal formulation encapsulating transcrocetin (TC) developed to enhance the diffusion of oxygen in the body. Here, we report the final results of the phase I/II clinical trial (NCT04378920; EUDRACT2020–001393–30) initiated to identify an optimal regimen and to assess the activity of TC in the context of acute respiratory distress syndrome (ARDS). More specifically, LEAF-4L6715 was developed to treat patients with ARDS due to severe SARS-CoV-2 infection who have a ratio of partial arterial pressure to inspired fraction of oxygen (PaO2/FiO2 ratio) <200 treated with artificial ventilation support in an intensive care unit.

A total of 37 patients were treated (across 6 dosing cohorts) with LEAF-4L6715 given as an intravenous infusion for over 90 minutes. The dose of LEAF-4L6715 was increased until the transaminase levels were elevated and 4 grade 3 events occurred among 8 patients. The recommended dosage was determined to be a fixed concentration of 300 mg administered every 12 hours. An improvement in the PaO2/FiO2 ratio and SOFA score was observed. The overall 28-day survival rate of 81%.

This study identified the recommended dose for LEAF 4L6715 and the dose-limiting toxicity and showed an overall favorable risk/benefit profile. These preliminary findings are promising for the activity of LEAF-4L6715 but will require confirmation in a randomized phase III trial.

## Introduction

Acute respiratory distress syndrome (ARDS) depicts various symptoms including pulmonary oedema which leads to oxygen diffusion impairment and acute hypoxemia. Pulmonary oedema arises from alveolar injury usually caused by an inflammatory/infectious process, as seen in severe COVID-19 and other septic conditions.^1,2^ The impaired diffusion of oxygen is characterized by a low ratio of partial arterial pressure of oxygen (PaO2) to fraction of inspired oxygen (FiO2) (PaO2/FiO2 ratio) resulting from the alteration of the alveolar-capillary barrier.^3^ The natural product crocetin, and in particular the trans isomer transcrocetin (TC), has demonstrated an ability to enhance systemic oxygen diffusion by kosmotropic effects.^4-8^ Nonetheless, the therapeutic effects of crocetins, including TC, are limited by their poor solubility and relatively short half-life, which limits their therapeutic effects and hence hampers their clinical development. The use of nanomedicine in the context of COVID-19 demonstrated the ability of this field to react to the situation and to provide novel carriers enabling to solve some of those major challenges (*i*.*e*. solubility, stability, improved pharmacokinetic properties).^9-11^

Herein, we report LEAF-4L6715 (LEAF4life Inc., Woburn MA, USA), a liposome formulation encapsulating TC. LEAF-4L6715 is a spherical unilamellar lipid bilayer liposomal nanoparticle with a loading capacity of approximately 115 g/mol of TC designed to improve TC’s pharmacokinetic (PK) properties by allowing the gradual release of TC into the circulation over time. The preclinical assessment of LEAF-4L6715 demonstrated that it caused an increased half-life of TC by ∼6-fold and an improved exposure (AUC_0-inf_) of liposome-released TC by ∼12-fold when compared to the same concentration of free TC. In a mouse model for ARDS sepsis, the addition of LEAF-4L6715 improved the survival of the treated mice compared to the active control group.^12^ These experiments, along with some animal safety studies, have provided the supporting evidence to launch this clinical study. The urgency and dire unmet need caused by the COVID-19 pandemic has fast-forwarded the clinical development of LEAF-4L6715 in this multicentric open-label phase 1/2 clinical trial. This study reports the results of the phase I/II clinical trial.

### Study Design

This study was designed to identify a safe and optimal dosing regimen for the clinical use of LEAF-4L6715 in terms of the dose and schedule based on both the PK profile and the clinical safety and efficacy results (NCT04378920; EUDRACT2020–001393–30). This clinical study was designed to be adaptive and accommodate any dose and schedule changes as needed when factors such as safety, activity, or PKs emerged. An independent data monitoring committee (IDMC) was organized to monitor the data and to assess the decisions regarding the conduct of the study; a study steering committee was also established for this study. The TC was manufactured by Chemspec-API (New Jersey, US) with cGMP compliance through a multistep total synthesis approach. The final LEAF-4L6715 drug product was synthesized by the encapsulation of free TC into a liposomal formulation by LEAF4life Inc. (Woburn MA, USA).

A starting dose of 2.5 mg/kg/day of LEAF-4L6715 was selected, which represents one-tenth of the presumed no observed adverse event level (NOAEL) that has been determined in preclinical studies.^12^ A duration of 90 minutes for the intravenous (IV) infusion was utilized after taking into consideration the established tolerable infusion duration of similar approved liposomal products. Based on previous results, the PK target therapeutic range of free TC to improve the diffusion of oxygen and glucose was defined as 4 to 49 μg/mL.^13^

The safety profile of LEAF-4L6715 was assessed based on the Common Terminology Criteria for Adverse Events (AEs) version 4.0. A dose-limiting toxicity was defined by the occurrence of a grade 3–4 AE in more than 30% of the patients treated at the same dose level. In addition to defining the preliminary clinical activity of LEAF-4L6715, an improvement in the PaO2/FiO2 ratio by ≥25% at 24 h was targeted as the primary efficacy criterion. This margin was defined based on the intra-patient variability of <20% for the PaO2/FiO2 ratio. The secondary efficacy criteria were assessed over time and included the PaO2/FiO2 ratio over time, the PaCO2, body position (prone versus supine), a positive expiratory pressure value, the concurrent use of cardiovascular supportive drug (noradrenaline), the sequential organ failure assessment (SOFA) scores, and the survival rate at 28 days.

Signed informed consent from each patient or his/her legal representative was obtained before any of the procedures began. To be enrolled in this study, patients had to be older than 18 years old, have ARDS with a PaO2/FiO2 ratio of less than 200, be receiving artificial ventilation support in an intensive care unit (ICU). and have a life expectancy of at least 24 h. Patients were required to have normal liver function as defined by the alanine aminotransferase (ALT), aspartate aminotransferase (AST), alkaline phosphate levels (AP) measuring less than 3 times the upper limit of the normal value for the institution, a platelet count above >100,000 cells/mm^3^, haemoglobin > 8 g/dL, and an absolute neutrophil count of >1000 cells/mm^3^. Any patient enrolled in another therapeutic clinical trial with the same endpoint was excluded, and pregnant or breastfeeding patients and patients who had haemoglobinopathy or a known hypersensitivity to crocetins, LEAF-4L6715, or any of its excipients were also excluded. Patients receiving extracorporeal membrane oxygenation were also excluded. All patients were expected to be followed for up to 90 days from the start of treatment. The first version of the protocol was approved by the ethical committee of CPP Sud Méditerannée III under application number 2020.04.08_20.03.27.77425 on April 8^th^, 2020. The French agency of medicine (Agence National de Sécurité du medicament et des produits de santé; ANSM) approved the use of LEAF-4L6715 for clinical study in April11^th^, 2020 (registration number MEDAECNAT-2020–03–00066).

### Pharmacokinetic analysis

Blood samples were collected from subjects to determine the serum concentration of total drug and free transcrocetin (TC) prior to infusion and at 1.5 (end of infusion), 2, 4, 8, 12, 24 hours after the start of infusion each day over 5 days. The concentrations of total LEAF-4L6715 and free TC were both measured in each sample by Liquid Chromatography – Mass Spectrometry (LC-MS) measurement, with the same approach using an automated robotic station. Samples were analyzed using an Ultra Hight Performance Liquid Chromatography (UHPLC) coupled to a triple quadrupole Shimadzu LC-MS 8030 mass spectrometer, with an electrospray ionization (ESI) source in the negative mode. Calibration and quality control samples were analyzed in the same series of injections for all fractions. The LC-MS analyses were performed on a C18 Kinetex column (50 mm × 2.1 mm, 2.6 μm, 100 A) as a stationary phase and mobile phase composition was 0.05 % formic acid in water (A) and acetonitrile (B), Multiple reaction monitoring (MRM) mode was used for quantitation. Ibuprofen was used as an internal standard, its MRM transition was 204.9 to 161.15. The median MRM transition for crocetin was 239.15 and ranged between 321.0 to 283.0.

The maximal and minimal concentrations (Cmax and Ctrough) were obtained from the measures of concentration. The area under the concentration-time curve from zero to the last quantifiable concentration (AUC0-t) was calculated using the linear trapezoidal rule, using actual elapsed time values. Others PK parameters were calculated by a non-compartmental analysis (NCA) performed with Pkanalix2020, Monolix suite (Lixoft®). The details of the pharmacokinetic modelling can be found in the **Supplementary Information**.

## Results

A total of 37 patients were enrolled in the trial (**Figure 1**). The baseline characteristics were in line with the profile of patients experiencing a severe form of COVID-19 disease treated in the ICU with artificial ventilation (**Table 1**). The median age was 69 years (ranging between 34 and 83), and the common past medical history of these patients included obesity (48.6%), morbid obesity (21.6%), hypertension (40.6%), and diabetes (16.2%). The diagnosis of COVID-19 was based on nasopharyngeal smear results and/or tracheal aspiration in 86.5% and 75.7% of cases, respectively. Baseline chest CT scans confirmed severe or critical lesions in 43.3% of patients. The median time between the admission to the intensive care unit (ICU) and the first dose of LEAF-4L6715 was shortened over time from 9.8 days in cohort 1 to 2.6 days in cohort 6.

**Table 1.**
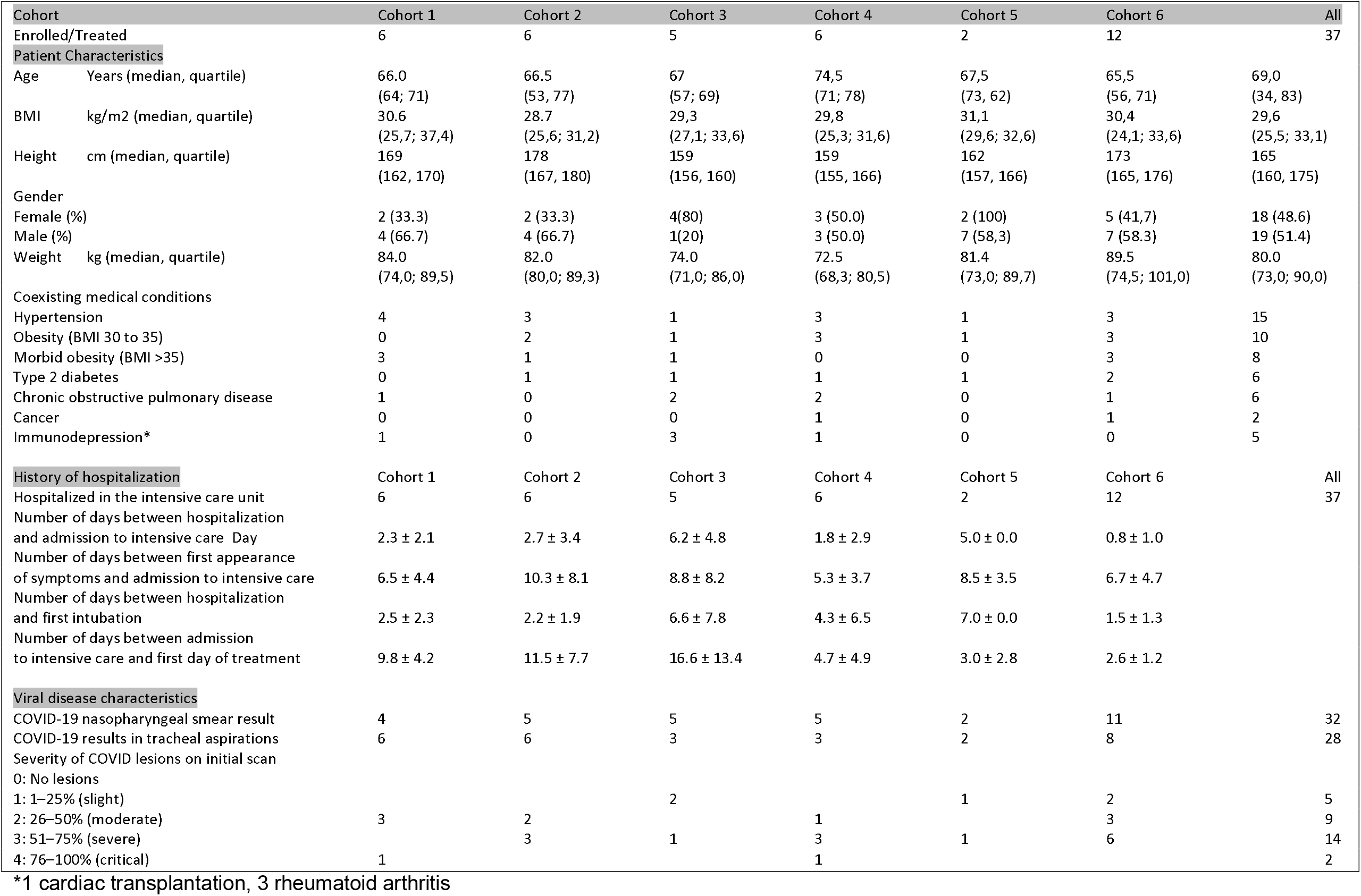
Patient characteristics

**Figure 1.**
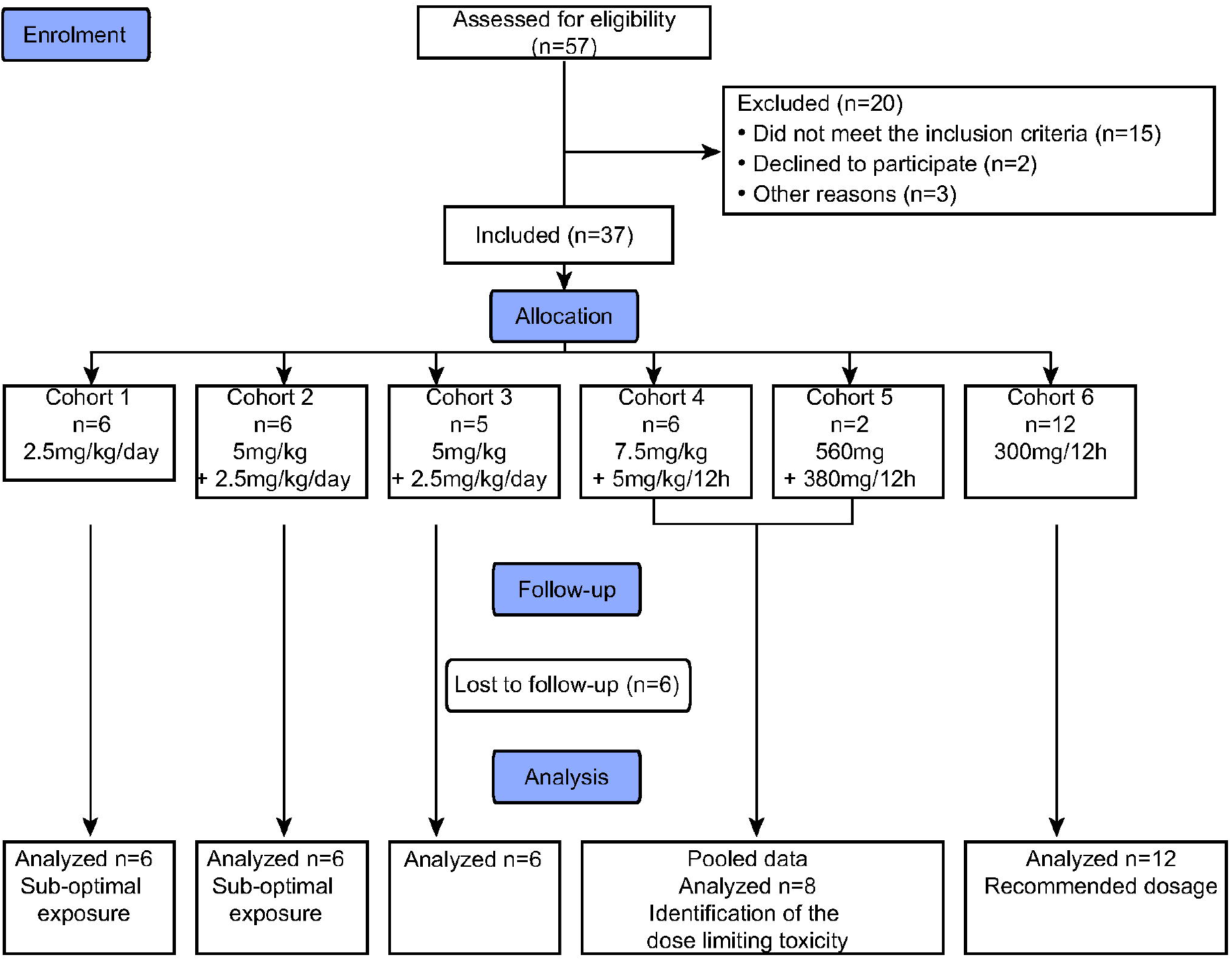
CONSORT flow diagram.

The clinical results from the patients enrolled in cohorts 1 and 2 have been preliminarily reported.^12^ Briefly, cohort 1 included 6 patients that were treated with LEAF-4L6715 at a dose of 2.5 mg/kg daily. No treatment-related serious AEs (SAEs) were observed. Nevertheless, the preliminary PK assessment of the total drug revealed the need to treat patients with LEAF-4L6715 for at least 3 consecutive days to reach a steady state. This prompted the addition of a loading dose in cohort 2. In cohort 2, 6 patients were subsequently treated with LEAF-4L6715 for over 30 cycles, with a loading dose of 5 mg/kg on day 1 followed by 2.5 mg/kg daily. One SAE reported was directly related to the treatment. Importantly, the results of the cohort 2 confirmed that thanks to the loading dose, the free drug concentrations reached the therapeutic range during the first 16 hours post-infusion. It was further noted that when following a maintenance dose of 2.5 mg/kg/day of LEAF-4L6715 starting on Day 2, the free TC level was inferior to the threshold of activity in 50% of the cases within 12 hours post infusion (the median of the free TC concentration was 0.58 μg/mL and ranged from 0 to 1.44 μg/mL).

In parallel with the enrolment in cohort 2, an extensive discussion emerged about the high proportion of patients with renal dysfunction who required renal replacement therapy by continuous venoveinous hemodiafiltration in ICUs and whether such patients should be included in the study. The IDMC recommended that the investigation of LEAF-4L6715 in patients with renal impairment should be undertaken, and a subset of patients with renal impairment were included in cohort 3. Of interest, the evaluation of the impact of renal impairment on the PK of LEAF-4L6715 was conducted by comparing the individual parameter estimates and the exposures attained in patients included in cohorts 2 and 3. The findings suggested that the impact of renal impairment on drug metabolism was limited and that dose adjustments based on renal function may not be warranted. Hence, based on these results from cohorts 1 and 2, a bicompartmental PK model was developed, suggesting that the administration of LEAF-4L6715 every 12 hours would be more appropriate to maintain a steady-state concentration of free TC within the prespecified therapeutic ranges.

These results supported the decision to increase the dosage in cohort 4 with a loading dose of 7.5 mg/kg, followed by a dose of 5 mg/kg with a shortened dosing interval of every 12 hours thereafter. Among the 6 patients enrolled in cohort 4, liver enzyme elevations were observed in four of the patients, with grade 3 elevations in 2 of these 4 patients. One of these grade 3 elevations, which was seen on day 5 was related to the administration of LEAF-4L6715 but reversible within 2 days after the end of treatment. The second grade 3 elevation was attributed to acute cholecystitis. The relationship with the study drug remains however unclear (**Table 2**). Noteworthy, the PK profiles confirmed that these patients were exposed to free TC within the prespecified targeted therapeutic range during the entire 24 hour period (**Table 3**).

**Table 2.** Safety profile of LEAF-4L6715

|  | Cohort 1 | Cohort 2 | Cohort 3 | Cohort 4 | Cohort 5 | Cohort 6 | All |
| --- | --- | --- | --- | --- | --- | --- | --- |
| N patients | 6 | 6 | 5 | 6 | 2 | 12 | 37 |
| N administration | 30 | 30 | 25 | 33 | 9 | 114 | 241 |
| Median administration<br>by patient (min-max) | 5<br>(5–5) | 5<br>(5–5) | 5<br>(5–5) | 5<br>(5–8)* | 5<br>(4–5)* | 5<br>(3–12)* |  |
| **Adverse Events (AE)** | 86 | 94 | 83 | 88 | 34 | 163 | 548 |
| Grade 1 | 43 | 43 | 36 | 42 | 17 | 88 | 269 |
| Grade 2 | 33 | 40 | 33 | 31 | 11 | 57 | 205 |
| Grade 3 | 9 | 10 | 14 | 7 | 5 | 16 | 61 |
| Grade 4 | 1 | 1 | 0 | 8 | 1 | 2 | 13 |
| Serious AE (SAE) | 3 | 2 | 0 | 8° | 2° | 5 | 19` |
| Death°° | 1 | 0 | 0 | 5 | 0 | 1 | 7 |
| AE leading to<br>treatment discontinuation | 0 | 0 | 0 | 0 | 1°°° | 1°°° | 2 |
| AE related<br>to study treatment | 12 | 7 | 8 | 9 | 7 | 27 | 70 |
| AE grade 3/4related<br>to study treatment | 2 | 2 | 0 | 3 | 1 | 4 | 12`` |
| SAE related to treatment | 0 | 0 | 0 | 0 | 1 | 0 | 1 |
| **AE of special interest (all grade)** |  |  |  |  |  |  |  |
| Cardiac |  |  |  |  | 1 |  | 1 |
| Skin | 1 | 1 |  |  |  |  | 2 |
| Renal | 6 | 4 | 9 | 7 | 0 | 4 | 30 |
| Digestif |  |  |  | 1 |  |  | 1 |
| Thoracic |  |  |  |  |  |  |  |
| Pneumopathy | 15 | 20 | 14 | 13 | 4 | 19 | 85 |
| Leucopenia | 0 | 0 | 0 | 1 | 0 | 4 | 5 |
| Thrombopenia | 0 | 0 | 1 | 1 | 0 | 0 | 2 |
| Anaemia | 7 | 14 | 8 | 12 | 3 | 11 | 55 |
| Hepatotoxicity | 10 | 5 | 8 | 6 | 6 | 23 | 58 |
| ASAT | 4 | 4 | 2 | 1 | 2 | 7 | 20 |
| ALAT | 4 | 1 | 0 | 1 | 2 | 6 | 14 |
| PAL | 2 | 0 | 6 | 4 | 2 | 10 | 24 |
| Billirubin | 6** | 4** | 3 | 3 | 1 | 5 | 22 |

\*administration every 12 hours \*\* artefactual assessment ° 4 grade 3 hepatic biology increase defining dose limiting toxicity °° None related to study treatment °°°
liver biology increase`2 haemorrhagic syndroms, 1 cholecystitis, 1 gas embolism, 5 deaths due to the aggravation of COVID-19 pneumopathy, and 10 liver biology increases.

**Table 3.** Pharmacokinetic parameters for LEAF-4L6715 regarding the total drug and free transcrocetine within each cohort

| Cohort | N | Dosing | Kel 24 h | R 24 h | T1/2 (h) | *C<sub>through</sub> | C<sub>max</sub> | AUC 0–24 h | V<sub>z</sub> | CI |
| --- | --- | --- | --- | --- | --- | --- | --- | --- | --- | --- |
|  |  | Scheme |  |  | =0,693/Kel | (µg/ml) | (µg/ml) | (h*µg/ml) | (L) | (L/h) |
| **TOTAL DRUG** |  |  |  |  |  |  |  |  |  |  |
| Median (IQR) |  |  |  |  |  |  |  |  |  |  |
| 1 | 6 | Every 24 h | 0,08<br>(0,07–0,08) | 9,92 | 9,1<br>(8,4–10,5) | 4,20<br>(3,4–5,8) | 28,25<br>(22,5–57) | 239,4<br>(194–367) | 9,06<br>(7,5–14,7) | 0,69<br>(0,49–0,88) |
| 2 | 6 | Every 24 h | 0,07<br>(0,06–0,08) | 0,88 | 9,9<br>(8,8–11,6) | 11,35<br>(9,7–16,2) | 72,90<br>(69,5–93,3) | 591,7<br>(506–708) | 7,05<br>(6,4–13,2) | 0,5<br>(0,44–0,59) |
| 3 | 4 | Every 24 h | 0,08<br>(0,08–0,10) | 0,99 | 9,2<br>(7,29–11,0) | 10,35<br>(8,6–15,5) | 84,05<br>(61,1–92,4) | 718,9<br>(614–832) | 5,82<br>(4,9–7,5) | 0,47<br>(0,39–0,55) |
| 4 | 6 | Every 12 h | 0,08<br>(0,05–0,11) | 0,84 | 8,3<br>(6,3–15,1) | 53,22<br>(38,5–57) | 120,4<br>(94,9–129,6) | 838,1<br>(744–1040) | 4,46<br>(3,6–5,1) | 0,37<br>(0,24–0,44) |
| 5 | 2 | Every 12 h | 0,05<br>(0,05–0,05) | 1 | 14,1<br>(14,1–14,1) | 65,15<br>(40,4–89,9) | 92,8<br>(89,9–95,7) | 1709,2<br>(721–2697) | 7,4<br>(7,4–7,4) | 0,36<br>(0,36–0,36) |
| 6 | 12 | Every 12 h | 0,06<br>(0,04–0,08) | 0,98 | 11,4<br>(8,9–17,3) | 22,35<br>(19,7–25,6) | 41,95<br>(34,9–45,9) | 330,8<br>(294–381) | 6,86<br>(6,2–7,9) | 0,44<br>(0,3–0,5) |
| **FREE DRUG (Transcrocetine)** |  |  |  |  |  |  |  |  |  |  |
| Median (IQR) |  |  |  |  |  |  |  |  |  |  |
| 1 | 6 | Every 24 h | 0,09<br>(0,08–0,1) | 0,62 | 7,70<br>(6,3–9,2) | 2,65<br>(0,3–3,9) | 21,75<br>(14,9–26,5) | 147,90<br>(112–185,1) | 14,72<br>(10,70–17,7) | 1,1<br>(0,8–1,9) |
| 2 | 6 | Every 24 h | 0,10<br>(0,07–0,11) | 0,83 | 6,90<br>(6,3–9,9) | 3,75<br>(1,5–8,2) | 46,10<br>(34,8–63,7) | 408,48<br>(265,3–408,5) | 15,08<br>(13,85–16,3) | 1,4<br>(0,9–2,3) |
| 3 | 4 | Every 24 h | 0,11<br>(0,06–0,15) | 0,77 | 6,30<br>(4,6–11,2) | 2,40<br>(0,9–6,0) | 24,95<br>(22,9–30,2) | 235,10<br>(220,9–255,3) | 13,27<br>(10,91–20,95) | 1,4<br>(1,2–1,7) |
| 4 | 6 | Every 12 h | 0,16<br>(0,1–0,23) | 0,88 | 4,33<br>(3,1–6,9) | 5,15<br>(4,5–6,8) | 20,15<br>(14,3–58,9) | 148,91<br>(107,4–282,7) | 27,88<br>(6,78–35,71) | 3,46<br>(1,6–3,9) |
| 5 | 2 | Every 12 h | 0,16<br>(0,16–0,16) | 0,92 | 4,3<br>(4,3–4,3) | 6,20<br>(2,9–9,5) | 12,15<br>(9,5–14,8) | 184,19<br>(83,4–285) | 34,19<br>(34,19–34,19) | 5,5<br>(5,5–5,5) |
| 6 | 12 | Every 12 h | 0,08<br>(0,08–0,09) | 0,87 | 8,25<br>(7,8–9,2) | 8,00<br>(6,9–8,9) | 15,50<br>(13,9–19,7) | 127,48<br>(99,2–147,5) | 16,51<br>(13,77–20,59) | 1,34<br>(1,1–1,7) |

\*the margins of biological activity ranged between 4 and 49 µg/ml for free transcrocetine

Based on the activity of LEAF-4L6715 in these cohorts, it was determined that a fixed dose regimen that is independent of patient weight may prove to be most practical, especially when considering the setting in which LEAF-4L6715 may have to be used most often, which would be in an ICU setting and for ARDS patients. The PK population modelling and simulation showed that a fixed dose given every 12 hours was comparable to a dosage based on the patient’s weight (**Supplementary information)**. Following these observations, in cohort 5, patients received a fixed loading dose of 520 mg followed by 380 mg every 12 hours; this represented the theoretical equivalent of the dose given in cohort 4. The PK results comparing cohorts 4 and 5 confirmed the validity of the fixed weight simulation, further supporting a fixed dose approach (**Table 3**). Interestingly, a cumulative effect for the total drug concentration was observed, and the need for the loading dosage to reach a steady state could be debatable. Nevertheless, the second patient enrolled in cohort 5 developed a grade 3 liver enzyme elevation on day 4, which required the discontinuation of treatment. The overall incidence of hepatic toxicity exceeded the dose-limiting toxicity threshold when cohorts 4 and 5 were combined and supported the assessment of a lower dose in Cohort 6 (at 4 mg/kg every 12 hours and without a loading dose). However, the robustness of the simulation supporting the fixed dose motivated the Steering Committee to propose a fixed dose of 300 mg, which is equivalent to 4 mg/kg for Cohort 6. Among the first 6 patients enrolled in this cohort, one patient experienced a transient grade 3 liver enzyme elevation. The PK profile confirmed that the free TC concentrations remained within the target therapeutic range during the entire treatment period. Cohort 6 was expanded to include six additional patients to confirm the previous findings. No additional grade 3–4 AEs were noted in this cohort.

### Safety results

Overall, 19 SAEs were reported in the study (**Table 2**), of which 10 were related to liver enzyme elevations, and 4 met the definition of the limiting toxicity for LEAF 4L6715 in cohorts 4 and 5. Two SAEs were haemorrhagic syndromes, one was cholecystitis, one was a gas embolism event linked to the manipulation of a central venous access, and 5 were deaths due to deterioration from COVID-19 pneumopathy. A total of 7 deaths occurred within the 30 days following the administration of LEAF-4L6715, and one occurred while the patient was receiving treatment beyond the initial 5 days period. None of the deaths were directly related to the experimental treatment, but instead were linked to the worsening effects of the pneumopathy. A total of 548 AEs were reported; the majority were not emergent treatments but were deemed to be consistent with the underlying disease and the ICU conditions. A total of 70 AEs were related to the treatment with LEAF-4L6715, of which 58 AEs were associated with liver enzyme increases. Among the AEs of special interest, no site infusion or acute hypersensitivity reaction was observed, and 1 cardiac event (atrial fibrillation) was reported. Additionally, the AEs of special interest included some grade III or IV anaemia events (16 patients), thrombosis and vascular disorders (6 patients), renal impairment (7 patients, of which 5 patients were in cohort 3), and respiratory, thoracic, and mediastinal disorders consistent with the underlying disease (16 patients), and all of the AEs were deemed to be unrelated to the study treatment.

The dose-limiting toxicity appeared to be liver toxicity, which was observed among 50% of patients (4/8) treated in cohorts 4 and 5, including 1 patient who experienced a grade III liver enzyme increase due to cholecystitis, which was retrospectively linked to treatment with LEAF-4L6715. In addition, 2 patients treated at this dose experienced grade 1 and 2 liver enzyme increases. In cohort 6, among the 12 patients taking the recommended dose, four grade 1 and 2 biological enzyme elevations were observed, and four grade 3 toxicity events were reported on day 5. One of the patients required treatment interruption; however, the elevations in the liver enzymes were found to be reversible, and all patients normalized to their baseline levels within a few days after the discontinuation of treatment. Of interest, all 12 patients treated in Cohorts 1 and 2 showed elevations in their total and free bilirubin levels (grades 1 and 2). After a careful investigation, we assessed whether the observed elevations in the levels of bilirubin were due to an artefact from the interference between LEAF-4L6715 and the assay used for the quantification of the bilirubin levels. The oxidation of TC induced a simultaneous absorbance of both bilirubin and TC at 450 nm, and the vanadate assay was used to measure bilirubin. On the other hand, when alternative methods were used for the assessment of bilirubin, including the diazonium salt, the 3,5-dichlorophenyldiazonium tetrafluoroborate (DPD)-based assay or the sulfanilic acid-based, the values of bilirubin fell within the normal range in the corresponding samples.

The most common toxicities observed were elevations in hepatic enzymes. After excluding the cases that had liver enzyme elevations that were attributed to artefactual bilirubin elevation, increased hepatic enzymes (at all grades) occurred in 65% of the patients (24/37) in the study.

### Pharmacokinetic analysis

The half-life ranged between 4–9 hours and 9–15 hours for free TC and total drug, respectively (**Table 3, Supplementary information**). The clearance and volume of distribution were comparable between all of the dosages for free TC and total drug. The AUC and C_max_ were proportional to the administered dosage, and no accumulation was observed for free TC over the 5 days.

### Therapeutic Activity of LEAF-4L6715

In cohort 6, which enrolled 12 patients who were treated at the recommended dosage, 25% (3/12) had an increase in the PaO_2_/FiO_2_ ratio >25% at 24 hours, and 67% of patients (8/12) had a >25% improvement at any time point during the treatment period (**Figure 2A-B, Table 4**). In cohort 6, within the 5 days of treatment, 25% (3/12) of the patients were extubated, a total of 6 patients were in the prone position at the start of treatment, and all patients but one were switched back to the supine position by day 5. The number of patients who required noradrenaline decreased from 6 patients (50%) at baseline to one by day 5. The SOFA, adjusted SOFA and cardiac SOFA scores significantly decreased over time (**Figure 2C**). Additional markers of improvement in respiratory function during treatment included a general decrease in the FiO_2_ requirement, a slight decrease in the PaCO_2_, a decrease in aggressive ventilation and a lower use of sedatives and paralytic agents.

**Table 4:** Activity criteria of LEAF 4L6715 in each cohorts

|  | Cohort # | Baseline | Day 1 (at 24hours) | Day 5 |
| --- | --- | --- | --- | --- |
| **PaO2/FiO2 ratio**<br>**(median [quartile])** | **1** | 148.1<br>(145.6-157.05) | 180.2<br>(162.5-208.6) | 115.3<br>(115.0-185.7) |
|  | **2** | 121.9<br>(118.4-126.8) | 168.0<br>(140.4-198.9) | 202.5<br>(192.3-236) |
|  | **3** | 148.1<br>(126.6-240.9) | 182.5<br>(126.6-282.4) | 192.1<br>(166.0-216.6) |
|  | **4+5** | 135.3<br>(118-162.2) | 140.3<br>(123.8-166.7) | 149.8<br>(120.8-172.5) |
|  | **6** | 129.1<br>(70.9-159.0) | 134.2<br>(122-154.8) | 160.0<br>(119.5-171.5) |
| **PEP (median [quartile])** | **1** | 12.0<br>(9.0-12.0) | 12.0<br>(9.0-12.0) | 10.5<br>(6.0-12.0) |
|  | **2** | 8.0<br>(7.0-9.8) | 10.0<br>(10.0-12.0) | 11.0<br>(10.0-12.5) |
|  | **3** | 6.0<br>(5.0-10.0) | 7.0<br>(7.0-9.0) | 8.0<br>(7.0-8.0) |
|  | **4+5** | 10.5<br>(8.5-11.8) | 11.0<br>(9.6-12.5) | 10.0<br>(9.0-10.0) |
|  | **6** | 10.0<br>(9.0-12.0) | 11.5<br>(9.5-13.5) | 12.0<br>(9.0-13.5) |
| **PaCO2 (median[quartile])** | **1** | 48.6<br>(46.4-50.2) | 45.0<br>(42.8-47.9) | 47.9<br>(45.4-51.0) |
|  | **2** | 41.7<br>(38.8-46.0) | 44.0<br>(39.5-51.8) | 44.5<br>(41.0-50.3) |
|  | **3** | 36.2<br>(35.1-41.0) | 42.8<br>(37.5-46.2) | 46.0<br>(36.9-46.6) |
|  | **4+5** | 37.5<br>(34.0-51.9) | 47.6<br>(43.1-53.7) | 50.3<br>(42.7-54.0) |
|  | **6** | 36.7<br>(34.1-41.7) | 40.7<br>(37.2-42.6) | 42.3<br>(37.3-44.4) |
| **Noradrenaline (%)** | **1** | 83,3 | 66,6 | 50,0 |
|  | **2** | 66,6 | 50,0 | 33,3 |
|  | **3** | 60,0 | 60,0 | 40,0 |
|  | **4+5** | 75,0 | 50,0 | 50,0 |
|  | **6** | 50,0 | 50,0 | 8.4 |
| **Position (%supine)** | **1** | 50,0 | 50,0 | 83,3 |
|  | **2** | 66,6 | 83,3 | 66,6 |
|  | **3** | 80,0 | 100,0 | 100,0 |
|  | **4+5** | 37,5 | 62,5 | 75,0 |
|  | **6** | 50,0 | 91,6 | 91,6 |

**Figure 2.**
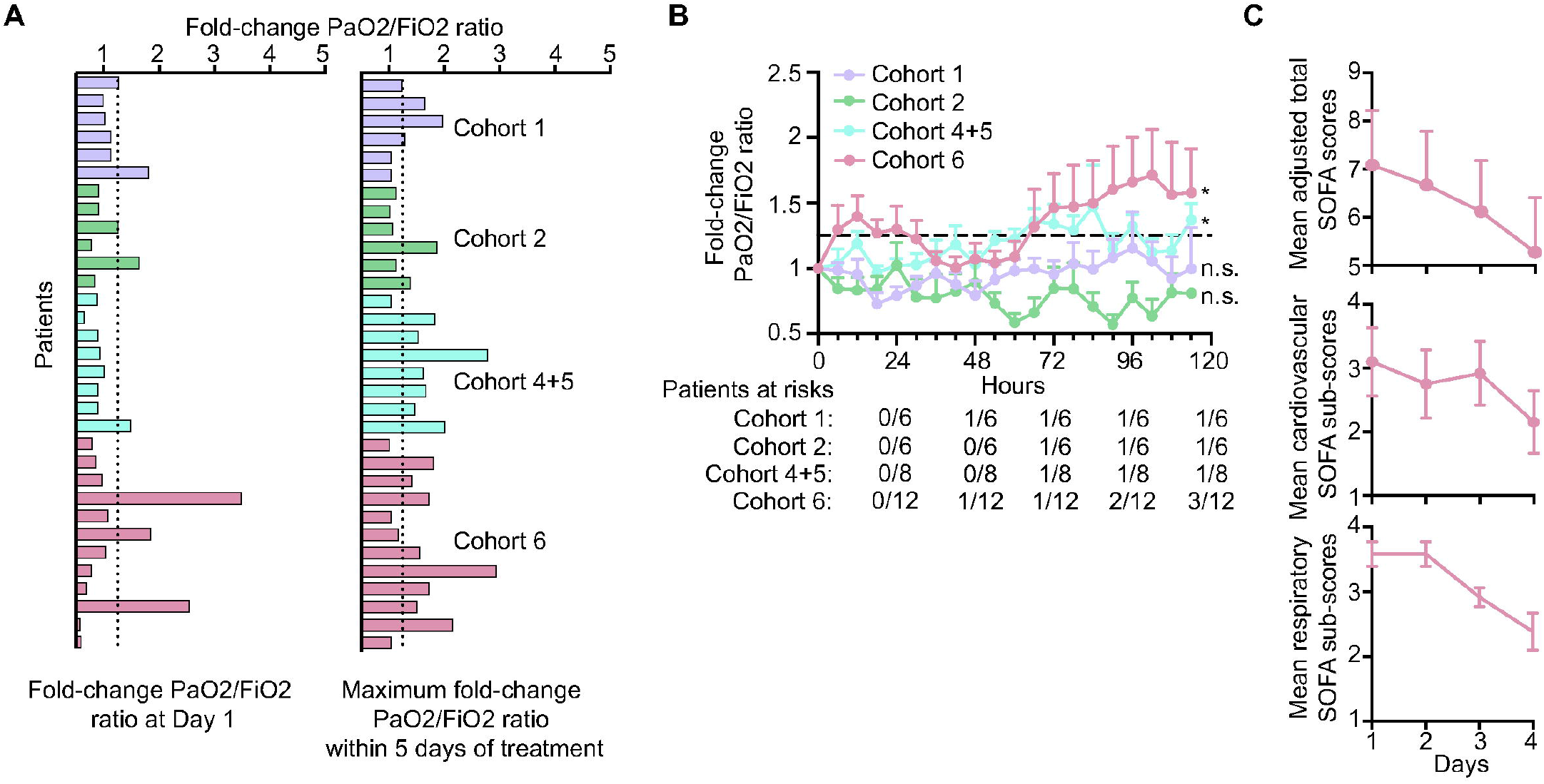
LEAF-4L6715 improves the reoxygenation in patients. **A**. PaO2/FiO2 ratio at 24 hours and at day 5 after initial treatment and **B**. within 5 days stratified by each cohort. **C**. SOFA score evolution of cohort 6.

A key secondary endpoint was the survival rate. Overall, 81% (30/37) of patients were alive at 28 days after enrolment. Subsequently 76% of patients were alive at 60 days and 73% at 90 days. Twenty five patients received 5 day cumulative doses ranging from 1000mg to 3000mg. Of these 24 (96%) remained alive at 28 days and 23 (92%) remained alive at 60 and 90 days of follow up. At the recommended dose of 300 mg every 12 hours (Cohort 6), 11/12 (91%) patients were alive at 28 days, at 60 and 90 days of follow-up.

## Discussion

ARDS has been identified as the primary cause of death in patients with diseases such as COVID-19 and sepsis. In the context of the COVID-19 pandemic, preliminary preclinical results have enabled us to trigger a fast-track clinical study to evaluate this novel liposomal drug.^12^

This study defined the recommended dose for the routine use of LEAF-4L6715, which was 300 mg of a fixed dose administered as an intravenous infusion over 90 minutes every 12 hours. Defining a fixed dosing regimen based on the PK modelling was a significant step in the development of this drug. Reversible liver enzyme elevations were identified as dose-limiting toxicities in this study. In addition, as many as 65% of the patients experienced all-grade hepatic toxicity in the study. This observation deserves further attention during the development of this new drug for three main reasons: i) because LEAF-4L6715 is a liposomal product, there is a potential for lipids to accumulate in the liver; ii) most intubated patients receive relatively high amounts of lipids parenterally; and iii) in this clinical setting, many patients present with underlying hepatic dysfunction. The combination of these three factors can lead to some potential hepatotoxicity, especially when one considers that high lipid levels in parenteral nutrition have been associated with hepatotoxicity in the ICU setting.

No other severe toxicity related to LEAF 4L6715 occurred in this study, and no additional safety concern appeared with the use of LEAF 4L6715 in this limited set of patients. The high incidence of AE occurrence seen in the study was considered to be due primarily to the underlying medical conditions and the associated care given in the ICU.^14^

From a PK standpoint, the small intracohort variability in the PK parameters suggested that the risk of drug interactions between LEAF-4L6715 and other concomitant medications used in the ICU is low. Comorbidities, including hepatic and renal disorders, had limited if any impact on the PK parameters, and this supports the use of a fixed dosing approach. The minimal interpatient variations observed for PK parameters in cohort 6 confirmed the robustness of the fixed dosing strategy for LEAF 4L6715. At the recommended dose of LEAF-4L6715, the exposure to free TC was always within the targeted therapeutic ranges in all patients during the entire course of treatment.

LEAF-4L6715 demonstrated encouraging activity as 25% of the patients treated at the recommended dose met the primary endpoint of a >25% improvement in the PaO2/FiO2 ratio at 24 hours, and up to 67% of patients showed a > 25% improvement in their PaO2/FiO2 ratio at any time point during the treatment period. Beyond this ratio, additional indicators of respiratory function, such as the requirement for FiO2, PaCO2, need for sedatives and paralytic agents, need for noradrenaline treatment, and number of patients requiring placement in a prone position, also showed improvement during treatment. Furthermore, the observed improvement in the total SOFA score and subscores indicates that treatment with LEAF-4L6715 may improve the microcirculation and thus the perfusion of other organs, thereby improving the function of many organs, in addition to the benefit observed in the respiratory system. Together, these observations point to the potential clinical benefit of LEAF-4L6715 in treating patients with COVID-19 and, more broadly, in treating patients with other diseases with a similar underlying pathophysiology, such as in sepsis, where multiorgan dysfunction is an important contributor to the morbidity and mortality. All these parameters of activity might reflect the treatment effect but require cautious interpretation because numerous factors may contribute to these results.

The observed 81% 28 day survival rate (76% at 60-days and 73% at 90 days) in this study appears to be promising. The therapeutic effect of TC is maximal within a certain range of concentrations, but if the concentrations are above or below, their therapeutic benefit diminishes. In a similar population, survival rates of 57% and 69% were observed following treatment with the standard of care both without and with dexamethasone, respectively (RECOVERY trial).^15^ Nevertheless, no prompt conclusion can be drawn because the survival rate varied widely in this population. Since the beginning of the COVID-19 pandemic, initial reports have raised concerns that the survival among patients receiving mechanical ventilation is exceedingly poor. Recently, evidence has arisen that the mortality rates might be more favourable and may be comparable to the mortality rates seen with ARDS when it is induced by other infectious pneumonias.^16,17^

Although the primary objective of the present study was to identify the optimal and safe dose and schedule of LEAF-4L6715, this study also generated promising efficacy data. However, the safety and efficacy findings from this clinical study will need to be confirmed in a randomized phase III study.

## Conclusion

This trial identified the recommended dose for treatment with LEAF-4L6715 as 300 mg given as a fixed dose, and LEAF-4L6715 is given over 90 minutes as an IV infusion every 12 hours. In this study, the dose-limiting toxicity was the elevation of liver enzymes, and there were no additional safety concerns. The overall risk/benefit profile of LEAF-4L6715 for the treatment of patients with ARDS due to COVID-19 who require artificial ventilation seems to be favourable. Furthermore, because hypoxia is an underlying risk factor for morbidity and mortality in many other disease states, LEAF-4L6715 should be investigated in other hypoxic medical conditions. The findings of this study, although promising, remain preliminary and need to be confirmed in a randomized trial. Meanwhile, given the urgent unmet need created by the ongoing COVID-19 pandemic, temporary authorization for use (ATU) was granted in France on November 17, 2020 for the use of LEAF-4L6715 in the treatment of patients with ARDS due to severe COVID-19.

## Data Availability

All data produced in the present study are available upon reasonable request to the authors

## Acknowledgements

The authors would like to acknowledge the ANSM (Agence Nationale de Sécurité du Médicament et des Produits de Santé) for its support towards the completion of the study.

## Authors’ Contributions

PMM, OC, PC, VC, VM, CN, VM, CB, PV, AD, and XP designed the study. The sponsor organized the clinical data collection (MV, AB). The PK study was carried out by PC, CB, HX, BG, and PV. Clinical data analyses were performed by ND, MV and AB. Data were interpreted by PMM, OC, XD, AR, PC, CN, VM, MCD, AD, and XP. AD and XP wrote the manuscript. All authors revised the manuscript and approved its submission. The corresponding authors have complete access to the data presented in the study and are responsible for submitting the manuscript.

## Data Sharing Statement

Data are available upon detailed and justified request to the sponsor of the study. The ANSM has previously reviewed all the presented data in this study and has full access to the data.

