## Supplementary Figures for "LEAF- 4L6715 enhances oxygenation in patients with acute respiratory distress syndrome (ARDS) due to severe COVID-19: Final results of a phase I/II clinical trial"

**Steady state achievement (single-dose vs. steady state pharmacokinetics) using population PK modeling.**

A) **PK parameters of total drug LEAF-4L6715**

The pharmacokinetics (PK) of LEAF-4L6715 has been evaluated in 18 patients with COVID disease. These patients were enrolled in 4 different cohorts. The first cohort included 6 patients that received 2.5 mg/kg of LEAF-4L6715 as an intravenous infusion of 90 minutes of duration once per day (QD) during 5 consecutive days. The second cohort enrolled 6 patients who received a loading dose of 5 mg/kg on Day 1 and 2.5 mg/kg of LEAF-4L6715 as an intravenous infusion of 90 minutes of duration QD on Days 2-5. The third cohort included 4 patients with renal impairment that followed the same dose regimen as those patients included in the second cohort. The last cohort included 2 patients who received a loading dose of 7.5 mg/kg on Day 1 twice per day (BID) and 5 mg/kg BID on Days 2-5 of LEAF-4L6715 as an intravenous infusion of 90 minutes of duration. On Day 5, the dose was reduced from 5 mg/kg BID to 2.5 mg/kg BID in one of these two patients included in cohort 4. Therefore, the PK of LEAF-4L6715 has not been evaluated in patients with COVID-19 following a single dose.

Using the data available of total LEAF-4L6715 concentrations, a population PK model was developed. A schematic representation of the population PK model developed is presented as below:

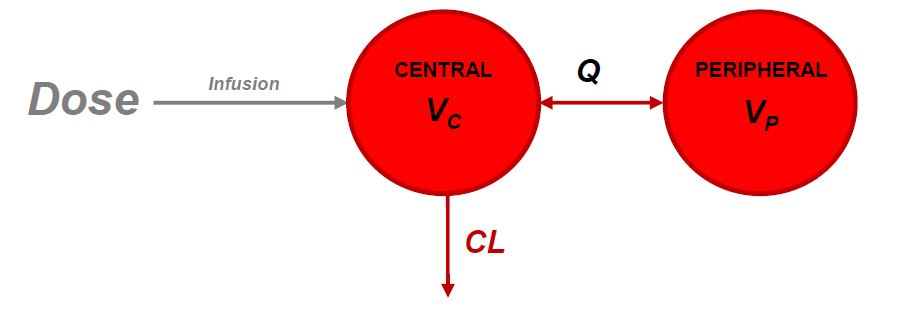

*Schematic representation of the population PK model developed to characterize total LEAF-4L6715 concentrations.*

In this model, CL, Vc, Q and Vp, represent the clearance, central volume of distribution, inter-compartmental clearance and peripheral distribution volume, respectively.

- CL was defined as:

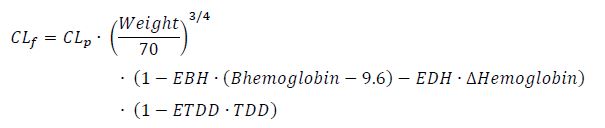

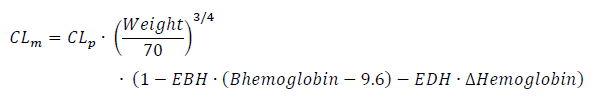

where CL_f_ and CL_m_ are the population CL for a female or a male, respectively. CL_p_ is the typical value of CL in the population. (Weight/70)^3/4^ is the effect of body size on CL (i.e. theory-based allometric exponent). Bhemoglobin is the baseline value of hemoglobin (i.e. the value of the hemoglobin at time 0). ΔHemoglobin is the individual difference at each time point in hemoglobin from baseline. EBH describes the effect of between-individual variation and corresponds to the fractional change in CL with each unit difference in Bhemoglobin from the median baseline hemoglobin (i.e. 9.6 g/dL). ΔHemoglobin describes the effect of hemoglobin variation within an individual and is the fractional change in CL with individual changes in hemoglobin. TDD is the total daily dose. ETDD is the fractional change in CL with each unit of increase in TDD. CL_p_, EBH, EDH, ETDD were estimated to be: 0.391 L/h/70 kg (for females) and 0.408 L/h/70 kg (for males), -0.201, -0.0985 and -0.0008, respectively. Therefore, the equations above may be written as follow:

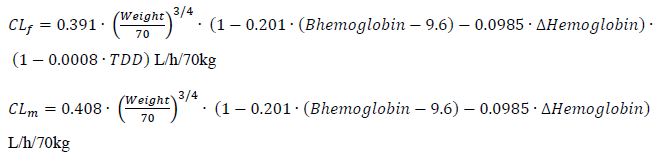

- Vc was defined as:

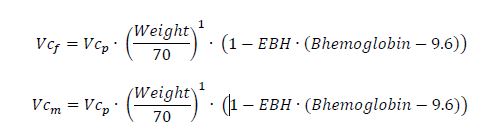

Where Vc_f_ and Vc_m_ are the population Vc for a female or a male, respectively. Vc_p_ is the typical value of Vc in the population. (Weight/70)^1^ is the effect of body size on Vc (i.e. theory-based allometric exponent). BHemoglobin is the baseline value of hemoglobine (i.e. the value of the hemoglobin at time 0). EBH describes the effect of between-individual variation and corresponds to the fractional change in Vc with each unit difference in BHemoglobin from the median baseline covariate (i.e. 9.6 g/dL). Vc_p_ and EBH were estimated to be 3.69 L/70 kg (for females) and 6.06 L/70 kg (for males), and -0.124, respectively. Therefore, the equations above may be expressed as:

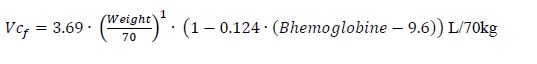

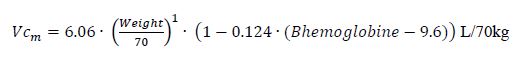

- Q was defined as:

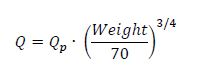

Where Q is the population Q. Q_p_ is the typical value of Q in the population. (Weight/70)^3/4^ is the effect of body size on Q (i.e. theory-based allometric exponent). Qp was estimated to be 0.767 L/h/70 kg and thus, the equation above can be written as:

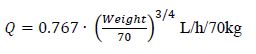

- Vp was defined as:

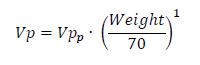

Where Vp is the population Vp. Vp_p_ is the typical value of Vp in the population. (Weight/70)^1^ is the effect of body size on Vp (i.e. theory-based allometric exponent). Vp_p_ was estimated to be 8.31 L/70 kg and thus, the equation above can be written as:

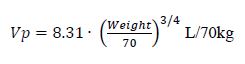

The population PK parameters of total LEAF-4L6715 are reported in Table S2. The goodness-of-fit (GOF) plots are depicted in Figure S3. Moreover, the visual predictive checks (VPC) is an internal validation tool that shows how well the model predicts the data on which the model was conditioned. It visualizes the median model prediction, the variability between patients (i.e. 5𝑡ℎ and 95𝑡ℎ percentiles), and the uncertainty around the predicted percentiles. The VPCs with the x-axis as time after the dose, time after the first dose, and stratified by cohort with the x-axis as time after dose, and time after the first dose are presented in Figure S3, Figure S4, Figure S5, and Figure S6, respectively.

Overall, the GOF plots of the population PK model of total LEAF-4L6715 did not present any major unexpected deficiency. Diagnostic plots showed tight normal scatter around the line of identity and showed an absence of significant bias. Furthermore, the distribution of the conditional weighted residuals did not indicate major deviation from the hypothesis of a symmetrical distribution.

**Table S2**. Population pharmacokinetic parameters of total LEAF-4L6715.

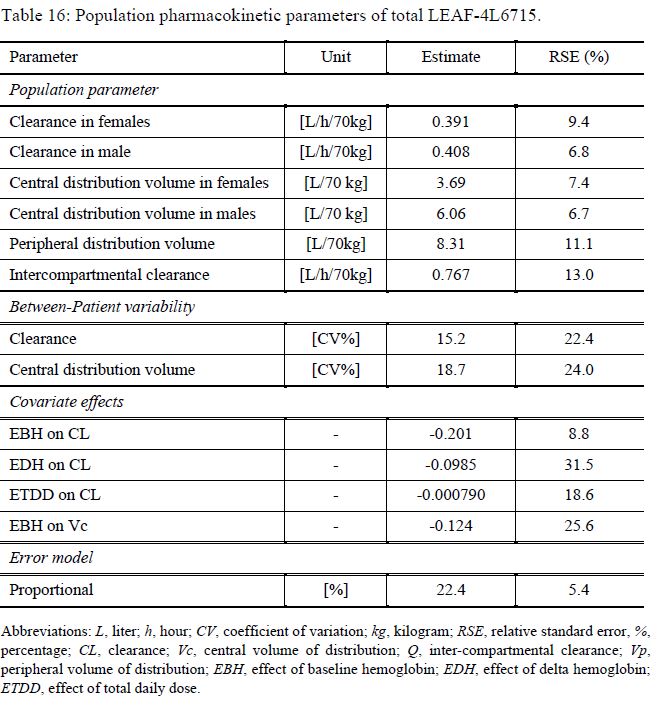

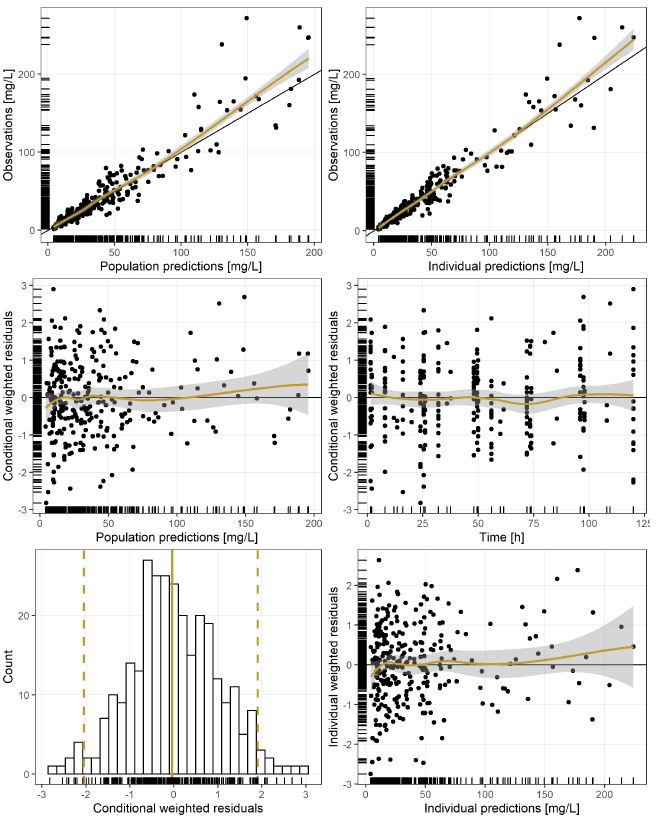
**Figure S3**. Goodness-of-fit plot of the population pharmacokinetic model developed to characterize the pharmacokinetics of total LEAF-4L6715 concentrations.

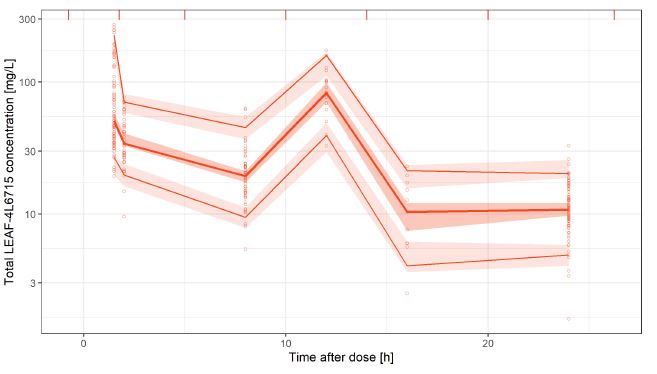

**Figure S4**. Visual predictive check with x-axis as time after dose.

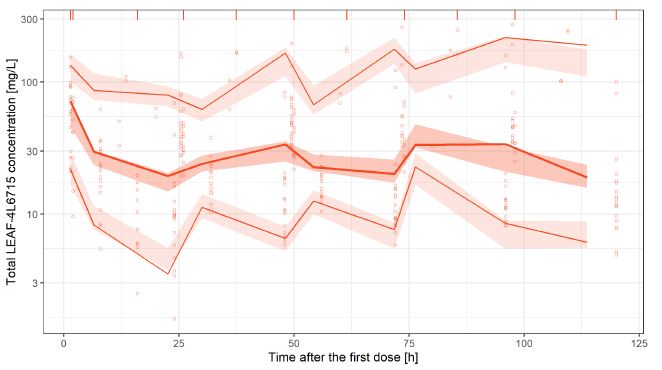

**Figure S5**. Visual predictive check with x-axis as time after first dose.

Overall, the data suggested that the proposed PK model was appropriate and sufficient to describe time course of total LEAF-4L6715 concentrations. This conclusion is supported by the VPCs because the observations (orange lines and points) were consistent with the simulated data (orange shaded areas).

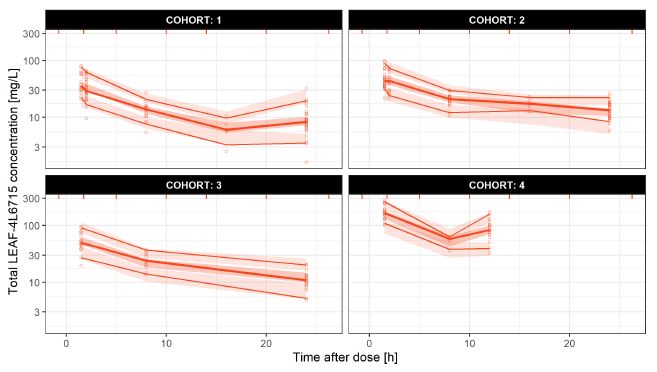

**Figure S6**. Visual predictive check with x-axis as time after dose stratified by cohort.

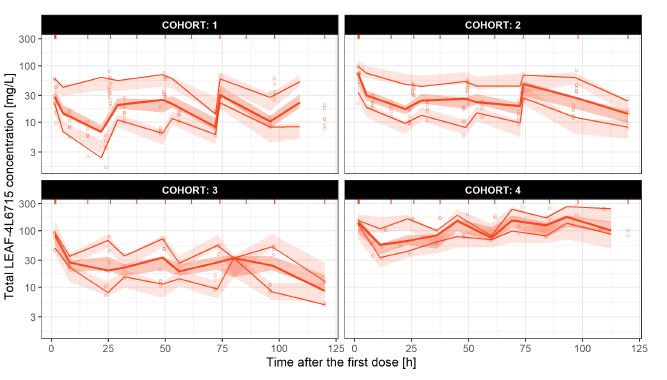

**Figure S7**. Visual predictive check with x-axis as time after the first dose stratified cohort.

Because the validation procedure was satisfactory, the model developed was used to predict exposure metrics. Table S3 summarizes the predicted minimum concentration (Cmin), maximum concentration (Cmax), and the area under the concentration-time curve (AUC) on Day 1 and Day 5 for each of the patients included in the analysis. Furthermore, Table S4 reports the mean values of the exposure metrics stratified by Day and cohort. Table S5 reports the key median population pharmacokinetic parameters of total LEAF-4L6715 stratified by cohort.

**Table S3.** Predicted exposure metrics on Days 1 and 5.

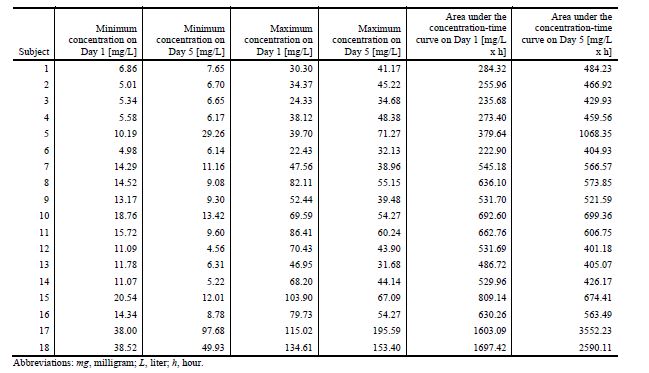

**Table S4.** Predicted mean exposure metrics on Days 1 and 5 stratified cohort.

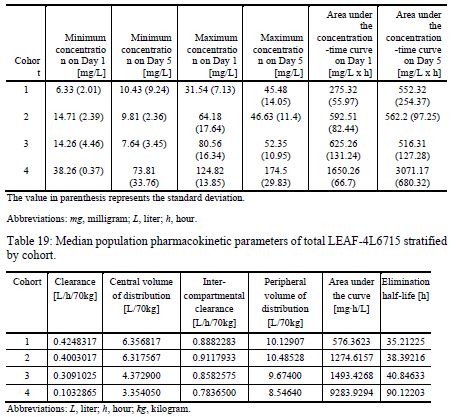

**Table S5.** Median population pharmacokinetic parameters of total LEAF-4L6715 stratified by cohort.

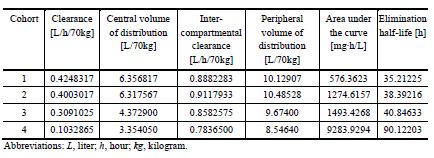

The model developed was also used to predict exposure metrics following single dose (Day 1) and multiple doses (Days 1-6). In order to predict the time course of total LEAF-4L6715 concentrations following a single dose, 1000 virtual females and 1000 males weighing 70 kg, with a baseline value of hemoglobin of 9.6 g/dL were assumed to receive a single dose of 200 mg of LEAF-4L6715 as intravenous infusion of 90 minutes of duration. Similarly, in order to simulate multiple doses, the same population described above was used to predict the time course of total LEAF-4L6715 concentrations. In this regard, 200 mg QD were assumed to be administered during 6 consecutive days. The exposure metrics were predicted on Day 6.

Table S6 reports the simulated exposure metrics following a single dose and following multiple doses. In addition, Figure S8 depicts the 95% prediction interval of the time course of total LEAF-4L6715 concentrations following a single dose (panel A) or multiple doses (panel B).

**Table S6**. Predicted exposure metrics following single and multiple doses in females and males weighing 70 kg, with a baseline value of hemoglobin of 9.6 g/dL receiving total LEAF-4L6715 as a single dose of 200 mg or as multiple doses of 200 mg during 6 consecutive days as an intravenous infusion of 90 minutes of duration.

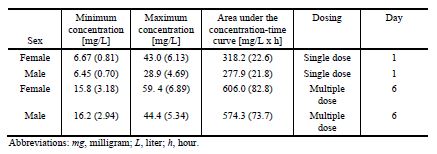

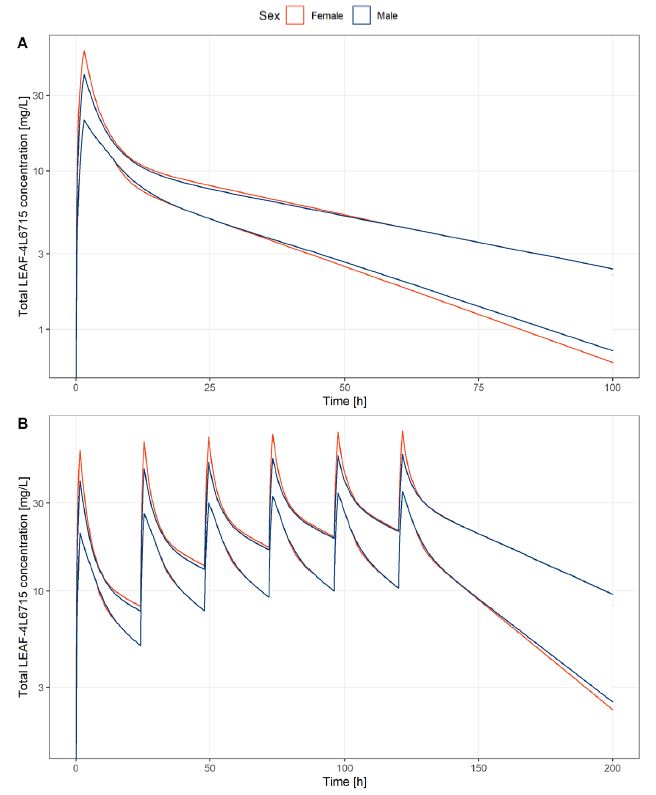

**Figure S8.** Predicted time course of total LEAF-4L6715 following a single dose (panel A) and multiple doses (panel B) and colored by females and males. The lines represent the 95% central prediction interval

***Summary of Exposure as Captured by Steady State Concentration***

The population PK analysis indicated that the PK of total LEAF-4L6715 concentrations was sufficiently characterized with a two-compartment model. The data obtained from the population PK model suggested that:

• The elimination half-life of total LEAF-4L6715 was 27 hours and 29.3 hours for females and males, respectively.

• As a general principle that it takes at least 5 half-lives to reach steady state concentration, therefore it is anticipated that it will take 6 days of treatment with LEAF-4L6715 given daily to achieve steady state concentrations.

B) **PK parameters of encapsulated and free LEAF-4L6715**

The schematic below represents the best population pharmacokinetic model to simultaneously characterize encapsulated and free LEAF-4L6715 concentrations

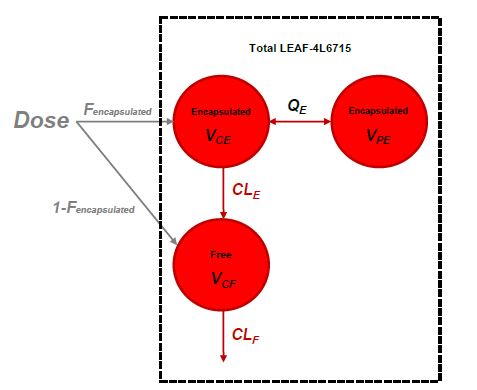

where CL_E_, V_CE_, Q_E_ and V_PE_, C_LF_ and V_CF_ represent the clearance, central volume of distribution, inter-compartmental clearance and peripheral distribution volume of the encapsulated, and clearance and central volume of distribution of the free trans-crocetin, respectively. F represents the fraction of total LEAF-4L6715 that is encapsulated immediately after drug administration.

- CL_E_ was defined as:

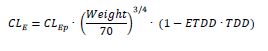

Where CL_E_ is the population CL_E_. CL_Ep_ is the typical value of CL_E_ in the population. (Weight/70)^3/4^ is the effect of body size on CL_E_ (i.e. theory-based allometric exponent). TDD represents the total daily dose administered and ETDD represents the fractional change in CLE for each unit of increase in TDD. CL_Ep_ and ETDD were estimated to be 0.405 L/h/70 kg and -0.0009, respectively. Therefore, the equation above can be written as follow:

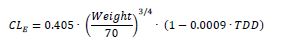

- V_CE_ was defined as:

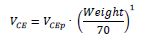

Where V_CE_ is the population V_CE_. V_CEp_ is the typical value of V_CE_ in the population. (Weight/70)^1^ is the effect of body size on V_CE_ (i.e. theory-based allometric exponent). V_Cep_ was estimated to be 1.16 L/70 kg. Therefore, the equation above can be written as follow:

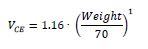

- Q was defined as:

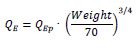

where Q_E_ is the population Q_E_. Q_Ep_ is the typical value of Q_E_ in the population. (Weight/70)^3/4^ is the effect of body size on Q_E_ (i.e. theory-based allometric exponent). Q_ep_ was estimated to be 1.67 L/h/70 kg and thus, the equation above can be written as:

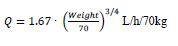

- V_PE_ was defined as:

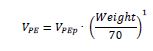

Where V_PE_ is the population V_PE_. V_PEp_ is the typical value of V_PE_ in the population. (Weight/70)^1^ is the effect of body size on V_PE_ (i.e. theory-based allometric exponent). V_Pep_ was estimated to be 7.58 L/70 kg. Therefore, the equation above can be written as follow:

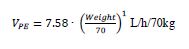

- C_LF_ was defined as:

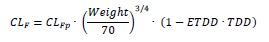

where C_LF_ is the population C_LF_. CL_Fp_ is the typical value of CL_F_ in the population. (Weight/70)^3/4^ is the effect of body size on CL_F_ (i.e. theory-based allometric exponent). TDD represents the total daily dose administered and ETDD represents the fractional change in CLF for each unit of increase in TDD. CL_Fp_ and ETDD were estimated to be 1.73 L/h/70 kg and -0.0006, respectively. Therefore, the equation above can be written as:

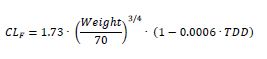

- V_CF_ was defined as:

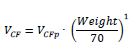

Where V_CF_ is the population V_CF_. V_CFp_ is the typical value of V_CF_ in the population. (Weight/70)^1^ is the effect of body size on V_CF_ (i.e. theory-based allometric exponent). V_CFp_ was estimated to be 3.32 L/70 kg. Therefore, the equation above can be written as follow:

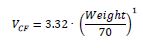

**Table S7**. Population pharmacokinetic parameters of encapsulated and free LEAF-4L6715 including all the patients with FE fixed to 85%.

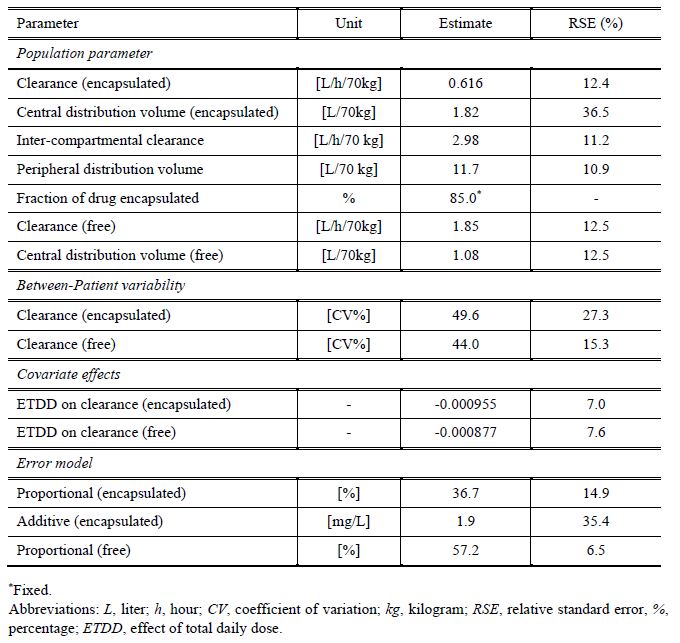

**Table S8.** Individual pharmacokinetic parameters of encapsulated and free LEAF-4L6715 including all the patients with FE fixed to 85%.

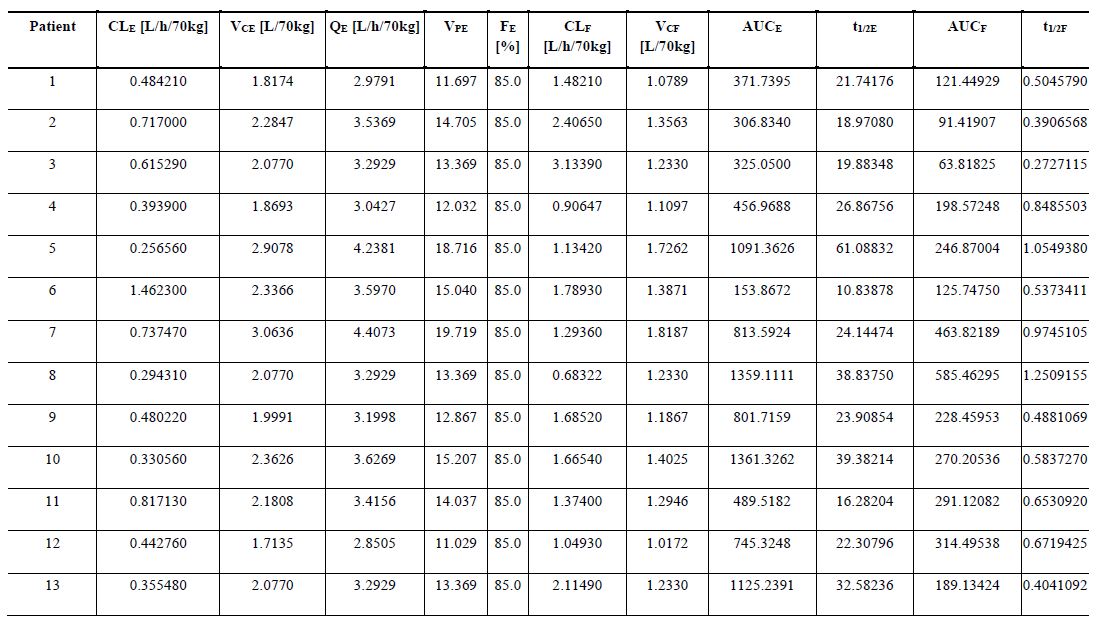

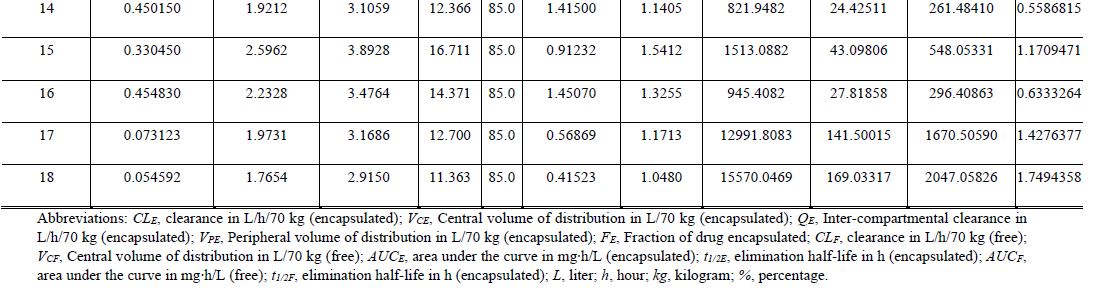

**Table S9.** Median population pharmacokinetic parameters of encapsulated and free LEAF-4L6715 including all the patients with FE fixed to 85% and stratified by cohort.

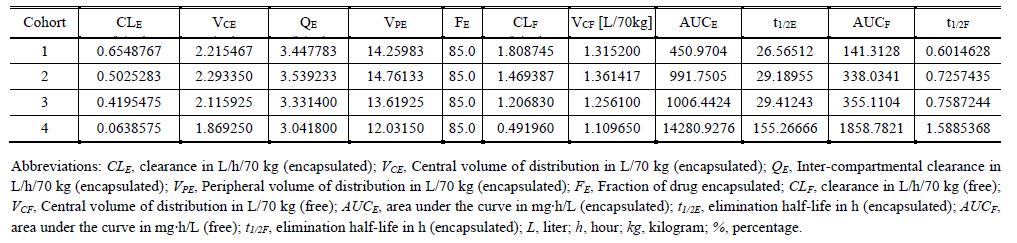
